# scRNA-seq revealed the special IG H&L V(D)J allelic inclusion rearrangement and the high proportion dual BCR expressing B cells

**DOI:** 10.1101/2023.04.13.23288510

**Authors:** Lanwei Zhu, Qi Peng, Yingjie Wu, Xinsheng Yao

## Abstract

Since the first report of V (D) J “ allelic exclusion/inclusion”and” dual BCR “in 1961, despite continuous new discoveries, the proportion and source mechanism of dual BCR under physiological conditions have been puzzling immuologists.This study takes advantage of the single cell V (D) J sequencing technology, which can perfectly match the heavy and light chains of BCR at the level of a single B cell, and obtain the full length mRNA sequence of the CDR3 region. By analyzing the pairing of functional IGH and IGL chains in single B cell from human and mouse bone marrow and peripheral blood, it was found that dual BCR B cells exhibit stable and high levels of expression.Among them, human bone marrow and peripheral blood contain about 10% dual (or multiple) BCR B cells, and mouse peripheral blood and bone marrow memory B cells contain about 20% dual (or multiple) BCR B cells. At the same time, we innovatively found that in each research sample of humans and mice, there are three (or more) functional rearrangements (mRNA level) of a single chain in a single B cell. By analyzing the position, direction and other compositional characteristics of the V(D)J gene family, we found that at least two(or more) of them are derived from over two(or more) specific allelic inclusion rearrangements of a single chromosome.(mRNA molecular level evidence).The results of this article provides new insights, new methods and modeling references for evaluating the proportion, molecular characteristics and source mechanisms of dual BCR B cells, as well as potential significance of allelic inclusion (exclusion escape) of V(D)J rearrangement.

## Introduction

The classical clonal selection theory of “one B cell-one antibody” [1] is the basis of B cell development, tolerance, and response, which relies on the V(D)J allelic exclusion rearrangement, the B cells did not conform to the rearrangement rules would undergo apoptosis, that is, the mature B cells express only one functional Heavy chain and Light chain. Allelic exclusion was proposed in 1957 and has been studied for nearly 60 years, but the detailed mechanism has not been clarified [2,3,4]. V(D)J allelic inclusion (incomplete allelic exclusion or allelic escape) rearrangement and dual BCR B cells have been widely supported by experiments since it was reported in 1961 [5]. However, due to the limitation numbers of single B cell studies, the proportion, mechanism and effects of dual BCR B cells on individuals still unclear [6,7,8,9]. Single cell sequencing (scRNA-seq) enables the analysis of a large number of single B cells with one (or more) Heavy chain and Light chain mRNA expressing, providing an opportunity for the intensive investigation of dual BCR proportion and mechanism. Xiaoyan Qiu et al analyzed the functional and non-functional recombination of two or more V-D-J (Heavy chain) and V-J (Light chain) recombination in single B cell of 5 healthy people by scRNA-seq [10]. Pelanda R et al compared the reorganization of dual K (Kappa-Light Chain, κ), dual L (Lambda-Light Chain, λ) and K+L in 3 healthy people and 3 SLE patients [11]. but they didn’t analyzed the B cell proportion and mechanism of the “H+K and/or H+ L” assembly “dual or multiple BCR”. According to the BCR rearrangement rule, even if both chromosomes are involved in rearrangement, a single B cell can only rearrange two types of Heavy chains, two types of K and two types of L chains [3,4,9]. We systematically analyzed human and mouse bone marrow and peripheral blood samples from multiple laboratories using published and shared “B cell single-cell sequencing V(D)J repertoire”, It is interesting to note that in the BCR sequence data of single B cells, we found three or more types of complete mRNAs of heavy or light chains in each sample [10, 11, 12, 13]. This raises the question of how the V(D)J allelic escape rearrangement occurs in these B cells.Meanwhile,for the first time, we found a high proportion of B cells expressing dual (or multiple) BCRs in single cell VDJ repertoire sequencing of human and mouse central and peripheral B cells (approximately 10% in human bone marrow and peripheral blood, approximately 20% in mouse memory B cells from peripheral blood and bone marrow).The detailed mechanism of formation and the biological significance of these B cells still require further research and exploration.

## Materials And Methods

1. The detail information of all samples The scRNA-seq datasets of B cells were download from 10X Company and Gene Expression Omnibus (GEO) data repository.Among them, human bone marrow sample (BM01) and peripheral blood sample (HB08),Mouse peripheral blood sample (PBMC_BALB/C,PBMC_C57BL/6) were all provided by 10X Company. Human peripheral blood samples (HB01, HB02, HB03) were healthy female volunteers aged 41, 36 and 68. (HB04,HB05,HB06)were all healthy volunteers aged 65 years. Mouse bone marrow samples (bm1,bm2):Single-cell suspensions from bone marrow of 3x NP-CGG immunized 8–12 weeks female C57BL/6 mice were prepared and CD19^+^ cells were enriched by magnetic cell sorting using anti-CD19 microbeads (Miltenyi Biotech). Ex vivo IgG1^+^/IgG2b^+^CD19^+^CD38^+^GL7^-^CD138^-^IgM^-^IgD^-^ memory B cells were isolated by FACS (Influx cell sorter (BD Bioscience)) and applied to the 10X Genomics platform using the Single Cell 5’ Library & Gel Bead Kit (10x Genomics) following the manufacturer’s instructions.
2. Definition of Functional and Non-functional B Cells. IGH (Functional) and IGL (Functional) sequences refer to in-frame recombination of functional V(D)J genes to form mRNA that can be completely transcribed, and then expressed as polypeptide chain sequences;Functional B cells refer to a single B cell contains at least one IGH (Functional) sequence and one IGL (Functional) sequence; non-functional B cells refers to the single B cells(1) no IGH (Functional) and IGL (Functional) sequences;(2) Only IGH (Functional) sequences were detected; (3) Only IGL (Functional) sequences were detected.
3. Functional H and L chain expression and combination proportion analysis process.
  1. One type of chain “cannot pair assemble BCR” B cells: H; K; L.
  2. Two types of chains “can pair and assemble into a single BCR”B cells: H+K;H+L;Two types of chains “cannot pair the assembled BCR” of B cells: H1+H2;K1+ K2;L1+ L2;K+ L.
  3. Three types of chains “can pair together into double BCR” B cells: H1+H2+K;H1+H2+L;H+K1+K2; H+L1+L2;H+K+L.Three types of chains “cannot pair the assembled BCR” B cells: H1+H2+H3;K1+ K2+ K3; L1+ L2+ L3;K1+ K2+ L;K+L1+ L2.
  4. Four types of (or more) chains “can pair B cells into three or more BCR”: H1+H2+H3+K;H1+H2+H3+L; H1+H2+K1+K2;H1+H2+L1+L2;H+K1+K2+K3;H+L1+L2+L3; Five or more are rare, and direct analysis does not show combinations: For example: H1+H2+H3+H4+H5, count to others.
4. Statistical analysis.

Data is presented as Mean ± SEM,statistical analysis with IBM SPSS Statistics 26 software was used to evaluate significant differences between the groups, and p<0.05 was considered statistically significant. GraphPad Prism software and Adobe Illustrator software were used for data visualization.

## Results And Discussion

We analyzed the scRNA-seq B cells BCR sequences of bone marrow and peripheral blood in the physiological condition of human and mice(Fig 1, Tab 1)The main results:

- H+K pairing was significantly higher than H+L in the single BCR B cells, and the proportion of mouse H+L pairing was much lower than that of human,the results consistent with the classical K superiority over Lambda utilization reported (Fig 1-B, Fig S2). And we found the dual (or multiple) BCR was mainly H+K+K, H+K+L, that is, K is also advantage utilization than L in dual BCR. In each sample, there are some H+H+n K/L and H+H+H+n K/L B cells (details examples in Fig 2). For the first time, a high proportion of dual (or multiple) BCR B cells (about 10% in human bone marrow and peripheral blood, about 20% in mouse peripheral blood and bone marrow memory B cells) were found (Fig 1-B,1-C, Fig S2), It were much higher than normal subjects (0.2-2%) [14,15] and normal mice (0.1-4%)[6,16] reported by FCM and other technical studies.
- A high proportion of single B cells containing three or more H (or K or L) chain V(D)JC mRNAs were found in both human and mouse samples. According to the position of a large number of V(D)J pairing families, V(D)J order on human and mouse H locus, direction of RSS, 12/23 deletional (looped out) and inversional rearrangement rules, and H chain D to J recombination preceding V to DJ recombination rules. We found the first time that three (or more) V(D)J mRNAs in a single B cell required a “single chromosome with specific allelic inclusion rearrangement”. BCR H chain required twice D-J inversional recombination on a single chromosome to form two types VDJC mRNAs transcripts (Fig 3-A,3-B: The rearrangement mechanism and examples). The K chain V-J required twice deletional or inversional rearrangements to form two types VJC mRNAs transcripts on a single chromosome (Fig S3). And the L chain V-J required twice deletional rearrangement to form two types VJC mRNAs transcripts on a single chromosome (Fig 3-C). If a single B cell can express four (or more) mRNAs only when both paternal and maternal chromosomes undergo twice (or more) rearrangements. Dual (or multiple) BCR B cells are mainly reported in autoimmune disease and plasmacytoma [8,9,11,17]. The RSS direction of V(D)J in IG gene locus is different from TR, both V and J of IGH can undergo deletional and inversional rearrangement with IGHD (V3’ with 7-23-9 RSS, D5’ and D3’ with 7-12-9 RSS, J5’ with 7-23-9 RSS), which greatly increases the accessibility of secondary or more rearrangements of the H chain in single chromosome. Individuals initiate rearrangements of alleles of the Heavy (or Light) chain of both paternal and maternal chromosomes, even twice and more rearrangement on one chromosome which has very positive implications for increasing the in-frame rearrangements of effective BCR and the diversity of BCR repertoire. However, the proportion, mechanism and significance of its occurrence remain to be studied in more depth by single-cell sequencing, such as the Heavy and Light chain pairing rule of dual BCR B cells, and functional studies at the protein level.
- In the process of analyzing the V(D)J sequences of all single B cell samples, we found that a certain proportion of B cells can only detect IGH (Functional) sequences or only IGL (Functional) sequences, and these cells can not express a functional BCR(Fig S1). and should not be included in the analysis of B cell outcomes. Therefore,our results suggest that the scRNA-seq of BCR should be analyzed separately by single BCR, dual or multiple BCR, and cells that cannot assemble BCR, so as to avoid bias or even errors of physiological or pathological significance in the analysis of single cell BCR repertoire.

**Figure 1.**
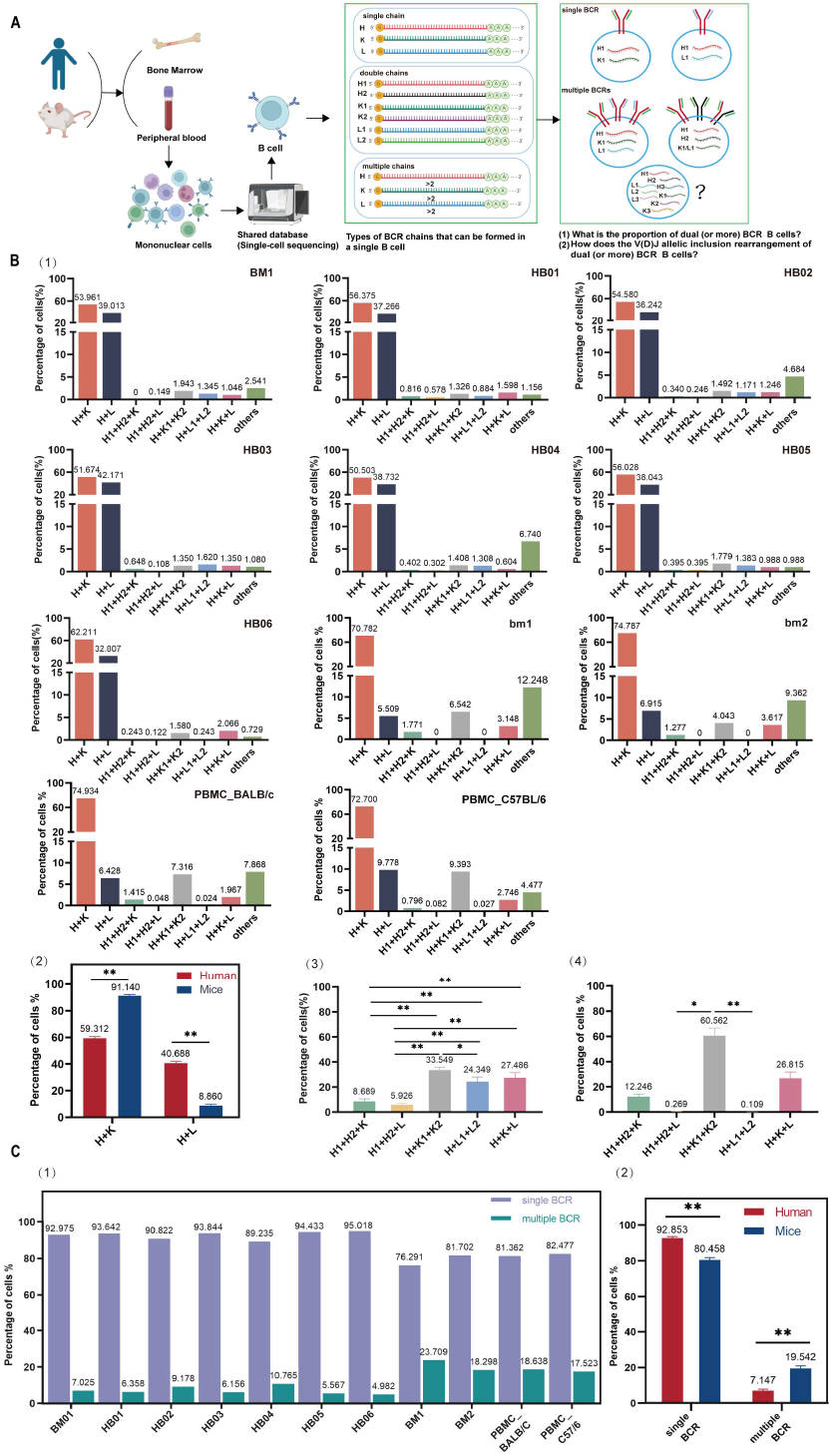
The high proportion dual (or multiple) BCR expressing B cells. **A**. Graphical abstract. Single-cell BCR sequencing samples from human and mouse bone marrow and peripheral blood were used to analyze the proportion and rearrangement mechanism of single BCR and multiple BCR **B (1)**. The proportion of Heavy and Light chains assembled into single and dual (or multiple) BCR expressing B cells in human and mice bone marrow and peripheral blood. **B(2)**. Statistical comparison of the proportions of H+K (mice>human) and H+L (human>mice) pairing of single B cells in human and mice. **B(3)**. Statistical comparison of the proportions of three chains (H, K, L) assembled into different types of dual BCR in human. The results was H+K1+K2>H+K+L>H+L1+L2>H+H+K>H+H+L. **B(4)**. Statistical comparison of the proportions of three chains (H, K, L) assembled into different types of dual BCR in mice. The results was H+K1+K2>H+K+L>H1+H2+K>H1 +H2+L>H+L1+L2. **C(1)**. The Proportion of single BCR and multiple BCR in human and mouse samples(single BCR>multiple BCR). **C(2)**. Statistical comparison of the proportions of single BCR and multiple BCR expressing B cells between human and mice. The proportions of multiple BCR in mice are higher than those of human multiple BCR. And it for the total number of sequenced cells **(Figure S2-B**) is slightly lower than it for the number of cells of the clone type in mice. **Note**. (1) The proportion was based on the number of cells of the clone type in each sample. In addition, the proportion (**Figure S2**) was based on the total number of sequenced cells for each sample. (2) Data in bar graphs are shown as mean+SEM. Statistical analysis was performed with unpaired t test, One-way Anova test,*p<0.05,**p<0.01.(3) H=Heavy Chain, K=Kappa-Light Chain, L= Lambda-Light Chain.

**Figure 2.**
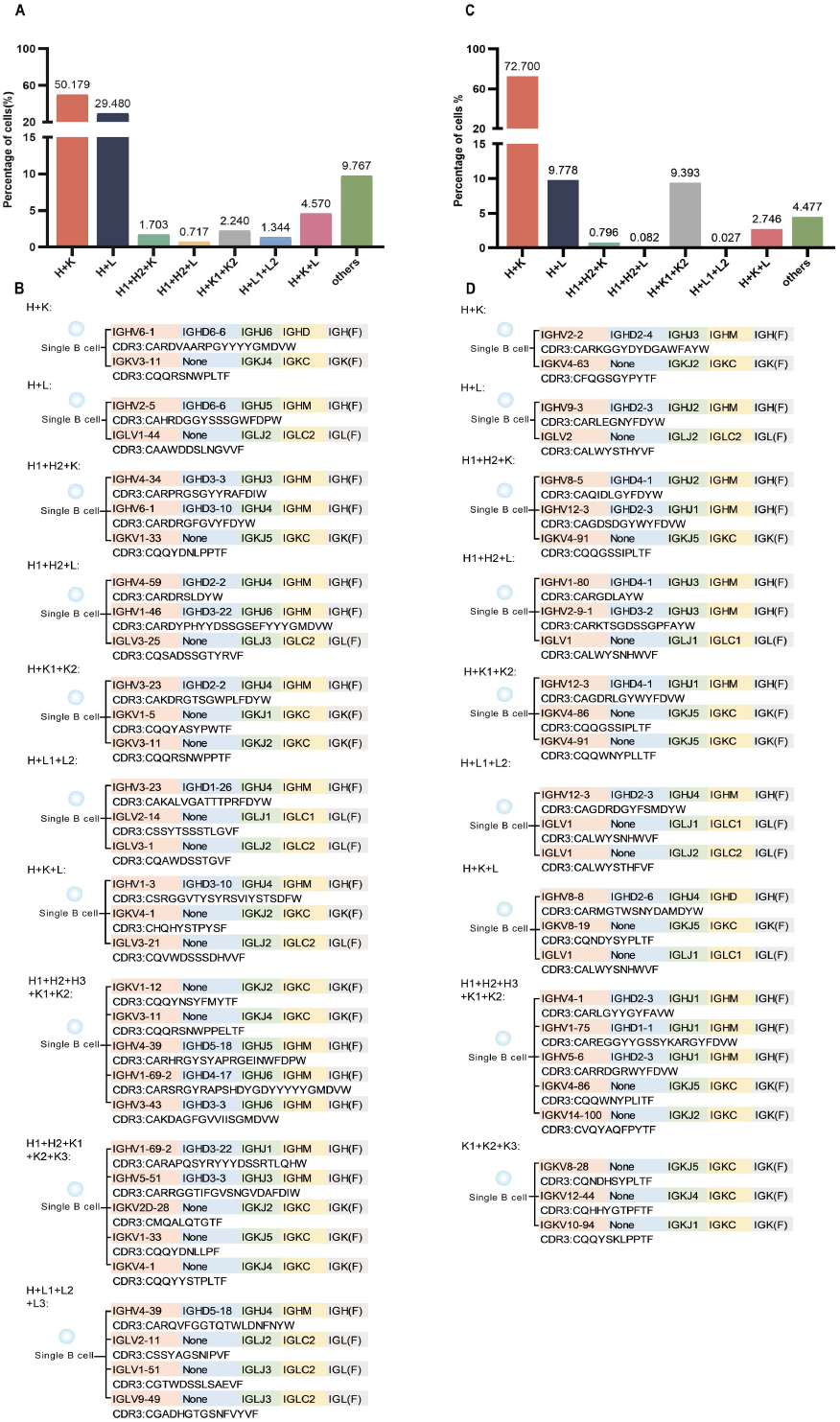
Examples of CDR3 sequences in a single B cell that can assemble into single BCR or multiple BCR in both human(HB08)and mouse(PBMC_C57BL/6). **A**. Pairing of Heavy and Light chains that can assemble into different types of single and dual (or multiple) BCR in human(HB08) sample. **B**. Examples of CDR3 sequences and the V(D)J recombination in a single B cell of human(HB08) sample. **C**. Pairing of Heavy and Light chains that can assemble into different types of single and dual (or multiple) BCR in mouse(PBMC_C57BL/6) sample. **D**.Examples of CDR3 sequences and the V(D)J recombination in a single B cell of mouse(PBMC_C57BL/6) sample. Note. The proportion of H+K pairing was significantly higher than H+L in the single BCR B cells of both human (HB08) and mouse (PBMC_C57BL/6) samples. However, the proportion of H+L pairing was much lower in mouse compared to human.Dual or multiple BCR in both species are mainly H+K+K and H+K+L, indicating that Kappa is utilized more frequently than Lambda in single or dual BCR. There are cells expressing 3 H chains (3 K chains or 3 L chains, etc) in “others”, examples of the VH(D)JH recombination patterns and CDR3 sequences in single cells expressing different types of single and dual (or multiple) BCR B cells in both human peripheral blood (HB08) and mouse (PBMC_C57BL/6) samples.

**Figure 3.**
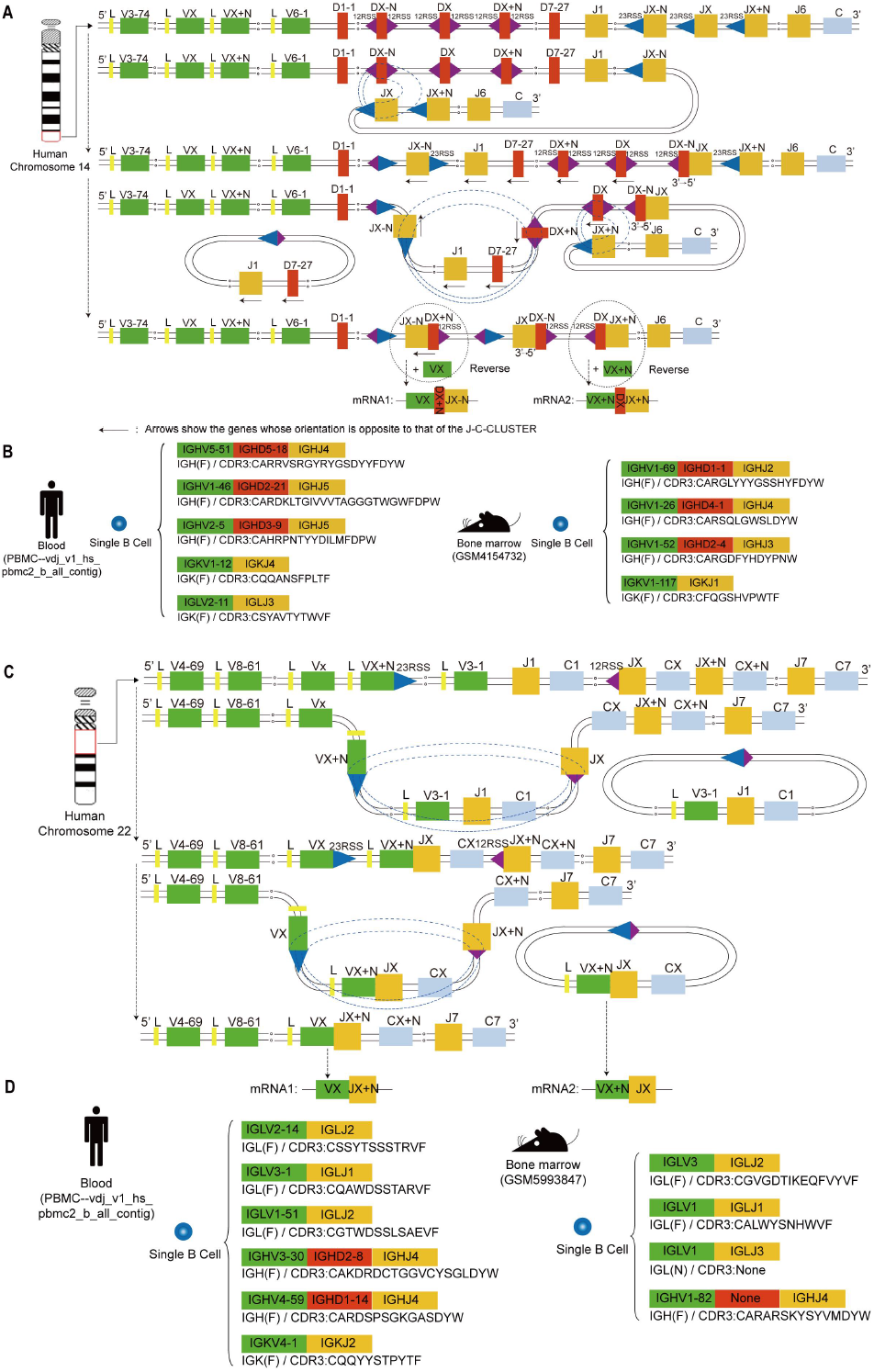
The Special rearrangement mechanisms for three types of IGH and IGL mRNA in a single B cell and an example of a single B cell CDR3 sequence in human and mouse samples. **A**. The H chain on a single human chromosome experiences two (or more) D-J inversional rearrangements. **B**. Examples of CDR3 sequences with three H chains from single B cells in both humans and mouse samples. **C**. The L chain on a single human chromosome experiences two (or more) V-J deletional rearrangements. **D**. Examples of CDR3 sequences with three L chains from single B cells in both human and mouse samples. **Note**. The H chain required two (or more) D-J inversional recombination on a single chromosome to form two (or more) VDJC mRNAs transcripts. (Among the three types of H in single human and mouse B cells as shown in the figure, at least two of them are derived from over two specific allelic inclusion rearrangements of a single chromosome).The L chain required twice deletional rearrangement to form two types VJC mRNAs transcripts on a single chromosome.(Among the three types of IGL in single human and mouse B cells as shown in the figure, at least two types of IGL are derived from specific allelic inclusion rearrangements in a single chromosome.)

**Table 1.** 13 samples basic information of single B cell BCR sequences of human and mouse bone marrow and peripheral blood and the proportion of B cells that can be assembled into single (or multiple) BCR

|  |  |  |  |  |  |  |  |  |  |  |  |  |  |
| --- | --- | --- | --- | --- | --- | --- | --- | --- | --- | --- | --- | --- | --- |
| Sample | Login number or<br>Filename | Total number of<br>primitive B cells | Total number of<br>analyzed B cells | The total number of<br>unique<br>sequences analyzed | Number of unique<br>clone type/total cells<br>that can be<br>assembled into BCR | H+K<br>(clone type/total<br>cells proportion%) | H+L<br>(clone type/total<br>cells%) | H1+H2+K<br>(clone type/total<br>cells %) | H1+H2+L<br>(clone type/total<br>cells %) | H+K1+K2<br>(clone type/total<br>cells %) | H+L1+L2<br>(clone type/total<br>cells %) | H+K+L<br>(clone type/total<br>cells %) | Others<br>(clone type/total<br>cells %) |
| BM01 | 10×Genomics | 1366 | 687 | 1424 | 669/673 | 53.961/54.086 | 39.013/38.930 | 0/0 | 0.149/0.149 | 1.943/1.932 | 1.345/1.337 | 1.046/1.040 | 2.541/2.526 |
| HB01 | GSM5550181 | 3439 | 3439 | 6709 | 2941/2970 | 56.375/56.263 | 37.266/37.273 | 0.816/0.808 | 0.578/0.606 | 1.326/1.313 | 0.884/0.909 | 1.598/1.684 | 1.156/1.145 |
| HB02 | GSM5550182 | 5431 | 5431 | 11476 | 5295/5307 | 54.580/54.626 | 36.242/36.216 | 0.340/0.339 | 0.246/0.245 | 1.492/1.489 | 1.171/1.168 | 1.246/1.244 | 4.680/4.673 |
| HB03 | GSM5550183 | 2189 | 2189 | 4238 | 1852/1889 | 51.674/51.509 | 42.171/42.456 | 0.648/0.635 | 0.108/0.106 | 1.350/1.323 | 1.620/1.588 | 1.350/1.323 | 1.080/1.059 |
| HB04 | GSM5831597 | 1120 | 1120 | 2970 | 994/1067 | 50.503/50.797 | 38.732/38.894 | 0.402/0.375 | 0.302/0.281 | 1.408/1.312 | 1.038/1.312 | 0.604/0.562 | 6.740/6.467 |
| HB05 | GSM5831600 | 1091 | 1091 | 2179 | 970/1012 | 56.392/56.028 | 38.041/38.043 | 0.412/0.395 | 0.412/0.395 | 1.546/1.779 | 1.237/1.383 | 0.928/0.988 | 1.031/0.988 |
| HB06 | GSM5831601 | 893 | 893 | 1793 | 823/843 | 62.211/62.278 | 32.807/32.266 | 0.243/0.237 | 0.122/0.119 | 1.580/1.779 | 0.243/0.237 | 2.066/2.135 | 0.729/0.949 |
| * HB07 | 10.1038/s41421-019-0137-3 | 5562 | 5561 | 11633 | /5180 | /50.598 | /36.544 | /0.560 | /0.290 | /1.409 | /1.795 | /5.251 | /3.552 |
| * HB08 | 10×Genomics | 3917 | 1202 | 2725 | 1111/1116 | 50.135/50.179 | 29.523/29.480 | 1.710/1.703 | 0.720/0.717 | 2.250/2.240 | 1.350/1.344 | 4.500/4.570 | 9.811/9.767 |
| average | / | / | / | / | / | 55.099/54.040 | 37.753/36.678 | 0.409/0.516 | 0.274/0.323 | 1.521/1.620 | 1.115/1.231 | 1.263/2.089 | 2.566/3.458 |
| bm1 | GSM4154732 | 3228 | 3224 | 7007 | 2033/2925 | 70.782/75.864 | 5.509/7.453 | 1.771/1.231 | 0/0 | 6.542/4.615 | 0/0 | 3.148/2.256 | 12.248/8.581 |
| bm2 | GSM4154733 | 1319 | 1317 | 2852 | 940/1225 | 74.787/75.184 | 6.915/10.531 | 1.277/0.980 | 0/0 | 4.043/3.184 | 0/0 | 3.617/3.020 | 9.362/7.102 |
| PBMC_BALB/C | 10×Genomics | 9204 | 4828 | 10417 | 4169/4267 | 74.934/74.830 | 6.428/6.914 | 1.415/1.383 | 0.048/0.047 | 7.316/7.148 | 0.024/0.023 | 1.967/1.945 | 7.868/7.710 |
| PBMC_C57BL/6 | 10×Genomics | 10148 | 4737 | 9508 | 3641/3771 | 72.700/73.296 | 9.778/9.706 | 0.796/0.769 | 0.082/0.080 | 9.393/9.122 | 0.027/0.027 | 2.746/2.678 | 4.477/4.322 |
| average | / | / | / | / | / | 73.301/74.793 | 7.157/8.651 | 1.315/1.019 | 0.033/0.032 | 6.823/6.017 | 0.013/0.012 | 2.870/2.475 | 8.488/6.929 |

**Note.** \* The clone type cells of HB07 could not be obtained. And HB08 come from 10× company could not include the age information etc. So statistical comparison of the proportions of clone type cells in Figure 1 was not included HB07 and HB08.
1. The detail information of all samples

## Conclusion

This study takes advantage of the single cell V (D) J sequencing technology, which can perfectly match the heavy and light chains of BCR at the level of a single B cell, and obtain the full length mRNA sequence of the CDR3 region.In the central and peripheral samples of humans and mice, the proportion and mechanism of B cells assembled into “dual or multiple BCR” by “H+K and/or H+L” were focused on analysis.For the first time, in the central and peripheral B cell repertoire of humans and mice, we found a high proportion and stable expression of dual (or multiple) BCR B cells, and both single and dual (multiple) BCR B cells showed the expression of K chain is superior to that of the L chain.Meanwhile,during the analysis process of this study,we innovatively found that in each sample, there are V(D)JC mRNA sequences containing three or more H (or K or L chains) in a single B cell, which are derived from single chromosome with specific allelic inclusion rearrangement (mRNA molecular level evidence).The results of this article provides new insights, new methods and modeling references for evaluating the proportion, molecular characteristics and source mechanisms of dual BCR B cells as well as potential significance of allelic inclusion (exclusion escape) of V(D)J rearrangement.

## Data Availability

All data produced in the present work are contained in the manuscript

## Acknowledgements

Thanks to 10X Company, Shi,Z, Peterson,J.N, Wang P and Riedel R for providing shared single cell sequencing sample data. This study was supported by the National Natural Science Foundation of China (82160279) and the Guizhou Provincial Hundred level Talent Fund [No.(2018)5637].

## Author information

### Authors and Affiliations

1. Department of Immunology, Center of Immunomolecular Engineering, Innovation & Practice Base for Graduate Students Education, Zunyi Medical UniversityZunyi, China.

### Contributions

Xinsheng Yao completed the experimental design and paper writing, Lanwei Zhu, Qi Peng, and Yingjie Wu completed the data analysis and chart making.

### Conflict of interest

The authors declare no competing interests.

### Ethics declarations

This study does not involve ethical requests or approvals.

### Data and materials availability

All data are available in the main text or the supplementary materials.

## Notes

### Competing Interest Statement

The authors have declared no competing interest.

### Funding Statement

This study was supported by the National Natural Science Foundation of China (82160279) and the Guizhou Province High-level Innovative Talent Fund[No.(2018)5637].

### Author Declarations

(1) Bone marrow of healthy volunteers (BM01), Bone--10k_BMMNC_5pv2_nextgem_intron_Multiplex_vdj_b_all_contig. (2) Peripheral blood of healthy volunteers 1(HB01), https://www.ncbi.nlm.nih.gov/geo/query/acc.cgi?acc=GSM5550181. (3) Peripheral blood of healthy volunteers 2(HB02), https://www.ncbi.nlm.nih.gov/geo/query/acc.cgi?acc=GSM5550182. (4)Peripheral blood of healthy volunteers 3(HB03), https://www.ncbi.nlm.nih.gov/geo/query/acc.cgi?acc=GSM5550183. (5)Peripheral blood of healthy volunteers 4(HB04), https://www.ncbi.nlm.nih.gov/geo/query/acc.cgi?acc=GSM5831597. (6)Peripheral blood of healthy volunteers 5(HB05),https://www.ncbi.nlm.nih.gov/geo/query/acc.cgi?acc=GSM5831600. (7)Peripheral blood of healthy volunteers 6(HB06),https://www.ncbi.nlm.nih.gov/geo/query/acc.cgi?acc=GSM5831601. (8)Peripheral blood of healthy volunteers 7(HB07), 10.1038/s41421-019-0137-3. (9)Peripheral blood of healthy volunteers 8(HB08), PBMC--vdj_v1_hs_pbmc2_b_all_contig. (10)Mouse bone marrow sample 1 (bm1), https://www.ncbi.nlm.nih.gov/geo/query/acc.cgi?acc=GSM4154732. (11)Mouse bone marrow sample 2 (bm2), https://www.ncbi.nlm.nih.gov/geo/query/acc.cgi?acc=GSM4154733. (12)Peripheral blood of the BALB/c mice (PBMC_BALB/C), PBMC--vdj_v1_mm_balbc_pbmc_b_all_contig. (13)The peripheral blood of the C57BL/6 mice (PBMC_C57BL/6), PBMC-vdj_v1_mm_c57bl6_pbmc_b_all_contig.

## References

1. Burnet, SFM. The clonal selection theory of acquired immunity. Nashville: Vanderbilt University Press, 1959.

2. Casellas R, Shih TAY, Kleinewietfeld M, Rakonjac J, Nemazee D, Rajewsky K, & Nussenzweig MC. Contribution of receptor editing to the antibody repertoire. science, 291(5508), 1541–1544(2001).

3. Vettermann C, Schlissel MS. Allelic exclusion of immunoglobulin genes: models and mechanisms. Immunological reviews, 237(1), 22–42(2010).

4. Outters P, Jaeger S, Zaarour N, Ferrier P. Long-range control of V (D) J recombination & allelic exclusion: modeling views. Advances in Immunology, 128, 363–413(2015).

5. Mäkelä O, Nossal GJV. Study of antibody-producing capacity of single cells by bacterial adherence and immobilization. J. Immunol. 87, 4 5 7–463 (1961).

6. Ten Boekel E, Melchers F, Rolink AG. Precursor B cells showing H chain allelic inclusion display allelic exclusion at the level of pre-B cell receptor surface expression. Immunity, 8(2), 199–207(1998).

7. Rezanka LJ, Kenny JJ, Longo DL. 2 BCR or NOT 2 BCR – Receptor dilution: a unique mechanism for preventing the development of holes in the protective B cell repertoire. Immunobiology, 210(10), 769–774 (2005).

8. Xu D. Dual surface immunoglobulin light-chain expression in B-cell lymphoproliferative disorders. Archives of pathology & laboratory medicine, 130(6), 853–856(2006).

9. Pelanda R. Dual immunoglobulin light chain B cells: Trojan horses of autoimmunity?. Current opinion in immunology, 27, 53–59 (2014).

10. Shi Z, Zhang Q, Yan H, Yang Y, Wang P, Zhang Y, Deng Z, Yu M, Zhou W, Wang Q, Yang X, Mo X, Zhang C, Huang J, Dai H, Sun B, Zhao Y, Zhang L, Yang YG, Qiu X. More than one antibody of individual B cells revealed by single-cell immune profiling. Cell discovery, 5(1), 1–13(2019).

11. Peterson JN, Boackle SA, Taitano SH, Sang A, Lang J, Kelly M, Rahkola JT, Miranda AM, Sheridan RM, Thurman JM, Rao VK, Torres RM, Pelanda R. Elevated Detection of Dual Antibody B Cells Identifies Lupus Patients With B Cell-Reactive VH4-34 Autoantibodies. Frontiers in immunology, 13(2022).

12. Wang P, Luo M, Zhou W, Jin X, Xu Z, Yan S, Li Y, Xu C, Cheng R, Huang Y, Lin X, Yao L, Nie H, Jiang Q. Global Characterization of Peripheral B Cells in Parkinson’s Disease by Single-Cell RNA and BCR Sequencing. Front Immunol, 13, 814239(2022).

13. Riedel R, Addo R, Ferreira-Gomes M, Heinz GA, Heinrich F, Kummer J, Greiff V, Schulz D, Klaeden C, Cornelis R, Menzel U, Kröger S, Stervbo U, Köhler R, Haftmann C, Kühnel S, Lehmann K, Maschmeyer P, McGrath M, Naundorf S, Hahne S, Sercan-Alp Ö, Siracusa F, Stefanowski J, Weber M, Westendorf K, Zimmermann J, Hauser AE, Reddy ST, Durek P, Chang HD, Mashreghi MF, Radbruch A. Discrete populations of isotype-switched memory B lymphocytes are maintained in murine spleen and bone marrow. Nat Commun, 11(1), 2570(2020).

14. Giachino C, Padovan E, Lanzavecchia A. κ^+^λ^+^dual receptor B cells are present in the human peripheral repertoire. The Journal of experimental medicine, 181(3), 1245–1250(1995).

15. Fraser LD, Zhao Y, Lutalo PM, D’Cruz DP, Cason J, Silva JS, Dunn-Walters DK, Nayar S, Cope AP, Spencer J. Immunoglobulin light chain allelic inclusion in systemic lupus erythematosus. European journal of immunology, 45(8), 2409–2419(2015).

16. Rezanka LJ, Kenny JJ, Longo DL. Dual isotype expressing B cells [κ^+^/λ^+^] arise during the ontogeny of B cells in the bone marrow of normal nontransgenic mice. Cellular immunology, 238(1), 38–48(2005).

17. Sang A., Danhorn T, Peterson JN, Rankin AL, O’Connor BP, Leach SM, Torres RM, Pelanda R. Innate and adaptive signals enhance differentiation and expansion of dual-antibody autoreactive B cells in lupus. Nature communications, 9(1), 1–14(2018).

