## Supplementary material for "scRNA-seq revealed the special IG H&L V(D)J allelic inclusion rearrangement and the high proportion dual BCR expressing B cells": FigureS1-S3

**Supplementary information**

**Supplementary Figure**

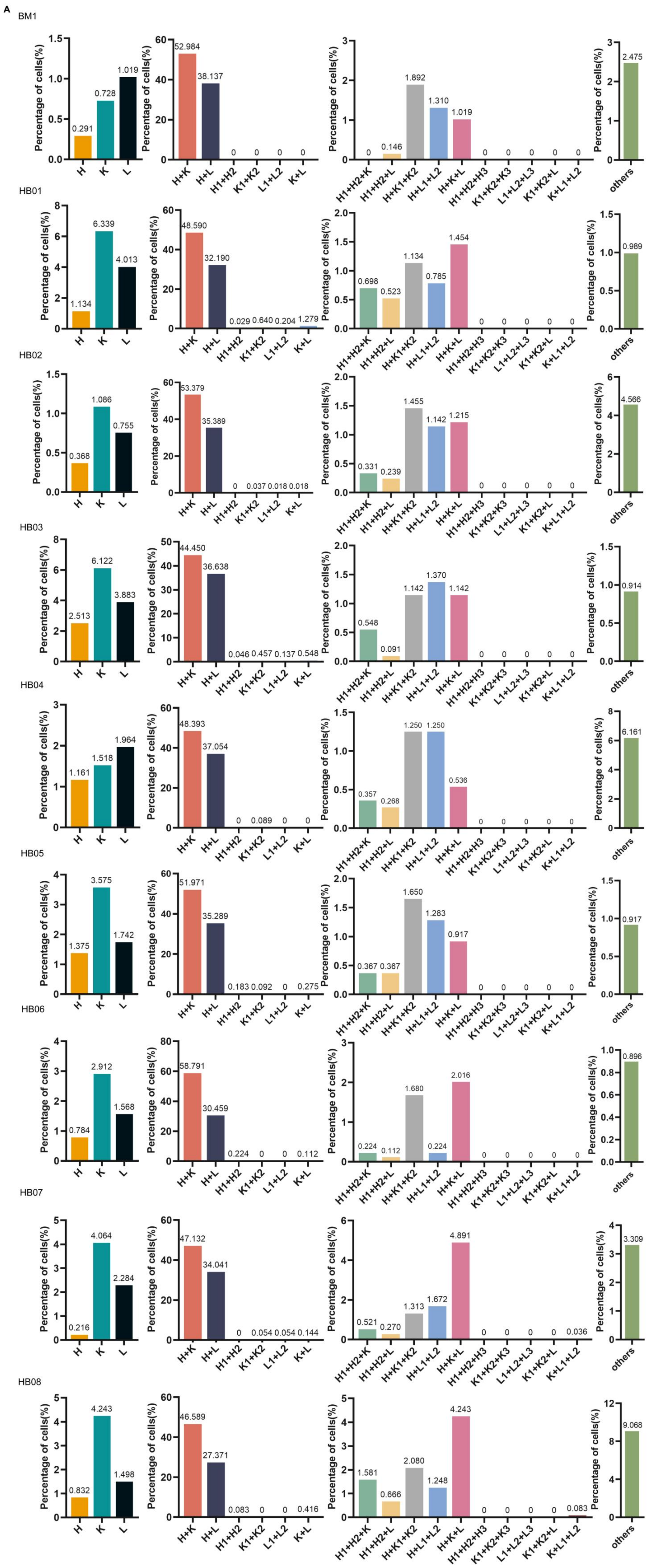

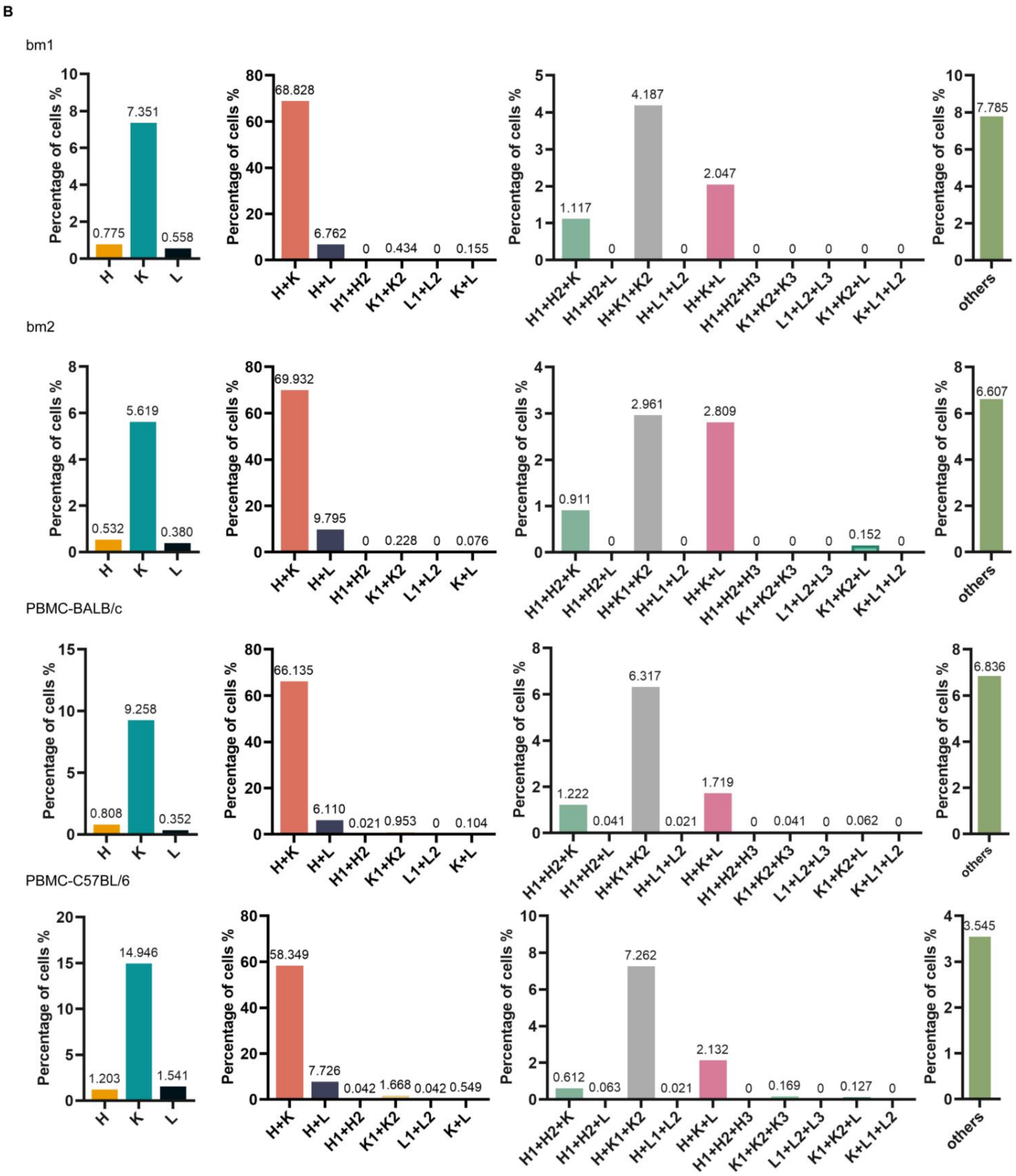


**Figure S1. The proportion of functional Heavy and Light chains in single B cell sequenced from bone marrow and peripheral blood of human and mouse.**

**A.** Human samples. BM1, HB01, HB02, HB03, HB04, HB05, HB06, HB07,HB08.

**B.** Mouse samples. bm1, bm2, PBMC-BALB/c, PBMC-C57BL/6.

**Note.** In each sample sequence of single cell sequencing BCR, there are sequences that functional Heavy and Light assemble into single BCR and multiple BCR, and sequences that functional Heavy and Light cannot assemble into BCR.

**Note.** (1) HB07 and H08 did not included in the Statistical analysis, because HB07 could not count the number of clone type cells. And HB08 did not included the age information. (2) The proportion of B cells in human bone marrow that can assemble single BCR (91.121%) was higher than that in peripheral blood (83.465%), the proportion of B cells that can assemble dual BCR (6.841%) was slightly lower than that in peripheral blood

(8.884%), and the proportion of cells that could not assemble BCR (2.038%) was lower than that in peripheral blood (7.650%). The proportion of memory B cells (77.695%) that assembled single BCR in bone marrow was slightly higher than that in peripheral blood (69.160%). The proportion of B cells that assembled dual BCR (14.212%) was similar to that in peripheral blood (14.833%). The proportion of cells that could not assemble BCR in bone marrow (8.130%) was lower than that in peripheral blood (16.007%). The proportion of single BCR cells in human (84.316%) was higher than that in mice (73.409%), and the proportion of dual BCR cells in mice and the proportion of cells that could not be assembled into BCR cells (14.522%, 12.069%) were higher than that in human (8.657%,7.027%).

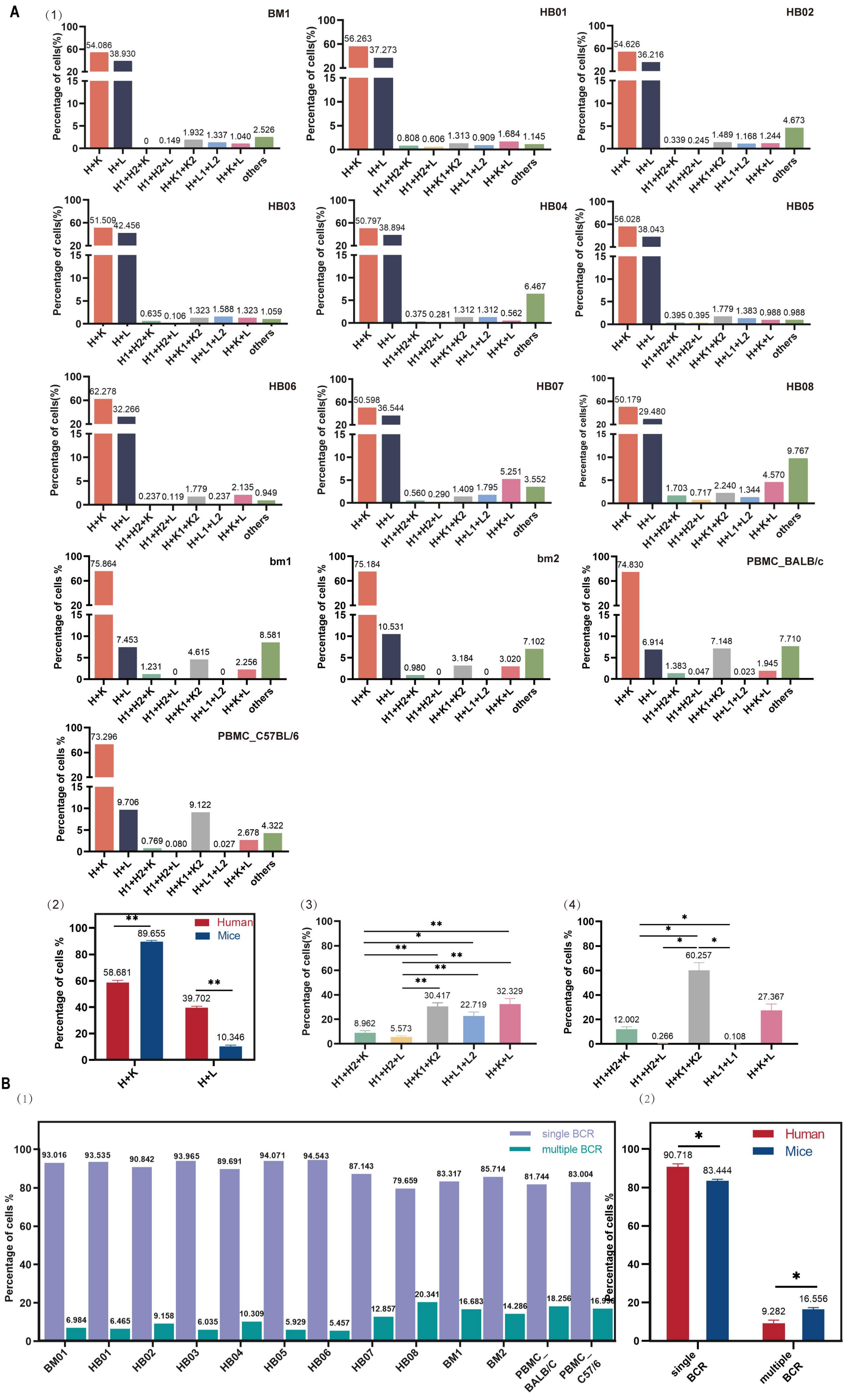

A
B
BM1
HB01
HB02
HB03
HB04
HB05
HB06
HB07
HB08
bm1
bm2
PBMC_BALB/c
PBMC_C57BL/6
Percentage of cells(%)
Percentage of cells %
H+K
H+L
H1+H2+K
H1+H2+L
H+K1+K2
H+L1+L2
H+K+L
others
Human
Mice
single BCR
multiple BCR

**Figure S2. The high proportion dual (or multiple) BCR expressing B cells**

**A(1)**.The proportion of Heavy and Light chains assembled into single and dual (or multiple) BCR expressing B cells in human and mouse bone marrow and peripheral blood.

**A(2)**.Statistical comparison of the proportions of H+K or H+L pairing of single B cells in human and mice. H+K pairing was significantly higher than H+L in the single BCR B cells, and the proportion of mouse H+L pairing was much lower than that of human.

**A(3)**.Statistical comparison of the proportions of dual (or multiple) BCR expressing B cells with different types of H and L chain pairings in human,The results was H+K+L>H+K1+K2> H+L1+L2>H1+H2+K> H1+H2+L.

**A(4)**.Statistical comparison of the proportions of dual (or multiple) BCR expressing B cells with different types of Heavy and Light chain pairings in mice,The results was H+K1+K2> H+K+L>H1+H2+K>H1 +H2+L>H+L1+L2.

**B(1)**.The Proportion of single BCR and multiple BCR between human and mouse samples（single BCR>multiple BCR）.

**B(2)**. Statistical comparison of the proportions of single BCR and multiple BCR expressing B cells in human and mice.（Except HB08, The proportions of multiple BCR in human were significantly lower than those of mouse multiple BCR）.

**Note**. (1)The proportion was based on the total number of sequenced cells for each sample. (2) Data in bar graphs are shown as mean+SEM. Statistical analysis was performed with unpaired t test, Mann-Whitney U test, One-way Anova test. **$p<0.05$,**$p<0.01$.

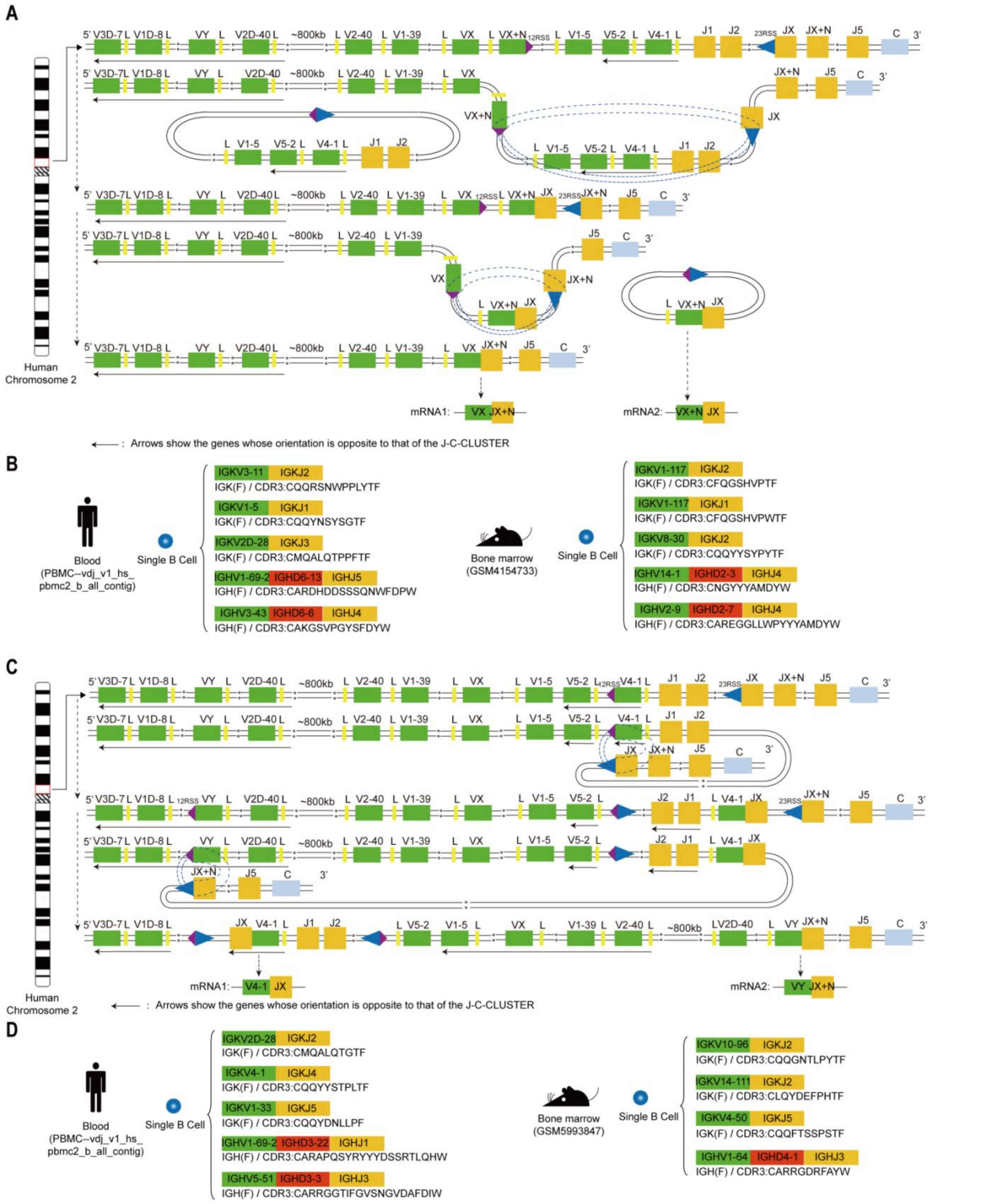


**Figure S3. The Special rearrangement mechanisms for three types of IGK mRNA in a single B cell and an example of a single B cell CDR3 sequence and V-J recombination in human and mouse samples.**

**A.** The K chain on a single human chromosome experiences twice (or more) V-J deletional rearrangements

**B.** Example of three V-J mRNAs in a single B cell by the mechanism of Fig S3-A.

**C.**The K chain on a single human chromosome experiences twice (or more) V-J inversional rearrangements

**D.** Example of three V-J mRNAs in a single B cell by the mechanism of Fig S3-C.

**Note.** The K chain V-J required twice deletional or inversional rearrangements on a single chromosome to form dual (or more) VJC mRNA transcripts. Among the three types of IGK in single human and mouse B cells as shown in the figure, at least two types of IGK derived from the specific allelic inclusion rearrangements in a single chromosome.
